# Incidence and determinants of antibiotic use by age two: a prospective cohort study

**DOI:** 10.64898/2026.09.23.26363799

**Authors:** Adrianna Staal, Harry Zhen, Toni Zhang, Morgan C. Byrd, Neil Kamdar, John R. Shaffer, Daniel W. McNeil, Mary L. Marazita, Betsy Foxman

## Abstract

**Background:** Half of US children are prescribed an antibiotic by age 2. Maternal education and socio-economic factors influence prescribing but the role of other factors, such as age of solid food introduction is unknown.

**Objectives:** We assessed risk factors for time to first antibiotic use between birth to age 2 to identify modifiable drivers of antibiotic use.

**Methods:** We analyzed interviews from mothers’ of 1032 children participating in the mother-child Center for Oral Health Research in Appalachia cohort 2 between 2011—2017. The cohort enrolled healthy, English-speaking, pregnant White women ≥18 years with singleton pregnancies. We used time-to-event analyses and a Cox Proportional Hazards model to describe and estimate hazard ratios (HRs) for time to first antibiotic.

**Results:** By age 2, 78% of children received at least one antibiotic (median age at first antibiotic: 12.5 months). Over half (58%) of antibiotics prescribed were for ear infections. After adjustment for household income and birth quarter as fixed variables, time of solid food introduction (aHR: 4.75; 95% CI: 3.19–7.07), time of first ear infection (aHR: 3.28; 95% CI: 2.58–4.17), and time of first illness (aHR: 4.60; 95% CI: 3.32–6.36) were significantly associated with time of first antibiotic use.

**Conclusions:** Earlier time of solid food introduction was associated with earlier antibiotic use. Childhood illnesses, especially ear infections, also strongly influence antibiotic initiation. These modifiable factors offer targets to reduce unnecessary early antibiotic use.

**Synopsis:** *Study Question:* What modifiable risk factors predict antibiotic use prior to age 2?

*What’s Already Known on This Subject:* Early childhood antibiotic use is common and influenced by clinical diagnoses like ear and respiratory infections. Socioeconomic factors shape prescribing patterns, but the contribution of early-life behaviors, including timing of solid food introduction, remains poorly understood.

*What This Study Adds:* This study identifies early solid food introduction as an independent predictor of earlier antibiotic use and quantifies the strong temporal links between childhood illnesses and initial antibiotic exposure, highlighting modifiable factors that may reduce unnecessary early antibiotic use.

## INTRODUCTION

Children under age two are prescribed antibiotics more frequently than any other age group in the United States, both in outpatient and ambulatory care settings.^1,2^ Over half of children receive at least one antibiotic prescription before the age of 24 months, with some estimates reaching as high as 70%.^3,4^ While antibiotics are essential for treating common bacterial pediatric illnesses like acute otitis media, tonsillitis, community-acquired pneumonia, pre-septal cellulitis, and urinary tract infections,^5^ concerns regarding early exposure and misuse remain significant. The widespread use of antibiotics among young children raises critical questions about their therapeutic necessity and potential long-term consequences, including microbiome disruption and increased risk of chronic health conditions.

Acute otitis media (AOM) is the most common ear infection in children, with five out of six children experiencing at least one episode before age three.^6^ In the U.S., the incidence rate of AOM in children aged 0–1 year is 66,283 per 100,000 person-years, and 94,589 per 100,000 person-years for children aged 1–2 years.^7^ In a New York cohort, 60% of children aged 6–36 months had at least one episode of AOM, with peak incidence between 6 and 12 months of age.^8^ Using U.S. National Ambulatory and National Hospital Ambulatory Medical Care surveys from 2006 to 2008, Hersh *et al*. estimated that 23% of the 49 million antibiotics prescribed to children less than 18 were for an AOM diagnosis, further emphasizing AOM’s impact on pediatric antibiotic use.^9^ However, it is estimated that AOM symptoms of six of every seven children with AOM will resolve without antibiotics.^10^ This, and concerns regarding increasing antibiotic resistance, led the American Academy of Pediatrics (AAP) to recommend watchful waiting prior to treatment for mild AOM.^11^

Inappropriate antibiotic prescribing increases the risk of multiple conditions in children and accelerates antibiotic resistance emergence and spread, leading to greater morbidity, mortality, and healthcare costs.^12,13^ Antibiotic exposure before the age of 24 months is associated with an increased risk of developing asthma, allergic rhinitis, atopic dermatitis, celiac disease, obesity, and attention deficit hyperactivity disorder.^3,14^ Two studies document the continued high rates of inappropriate antibiotic prescribing to children. Butler *et al*. tracked insurance claims data from diagnosis through follow-up visits and prescription records between 2016 to 2018 among 2.8 million children in the United States aged 6 months to 17 years.^15^ Thirty-six percent received inappropriate antibiotic prescriptions for bacterial infections (non-first-line antibiotics per target guidelines) and up to 70% received antibiotic prescriptions for viral infections where antibiotics are not indicated. Further, almost half (48%) of the children with non-suppurative otitis media – where watchful waiting and pain management are recommended^16^ – were prescribed antibiotics. Lehrer *et al*. evaluated the 506,633 antibiotic prescriptions prescribed in 2022 to Tennessee children aged <20 years, revealing that only 31.4% of antibiotics prescribed in outpatient settings were optimal in terms of choice and duration.^17^

Most otitis media episodes occur as a complication of upper respiratory infection.^18^ The prevalence of viral infection is very high in young children. In a study of 26 households sampled weekly for 52 weeks and tested for respiratory virus, children less than 5 tested positive in half the samples.^19^ Good hand hygiene can prevent spread of respiratory infections by 21%.^20^ Early introduction of solid foods, which increases exposure to caregivers hands and feeding utensils, may increase infant exposure to additional microbes beyond^21,22^ those expected from the increased mouthing behaviors that occurs during normal development.^23^ For example, in a laboratory-based simulation, Cho *et al*. demonstrated that caregivers’ hands and feeding utensils (e.g., spoons) act as reservoirs for pathogens like *Cronobacter sakazakii, Salmonella enterica*, and *Staphylococcus aureus* when feeding powdered infant formula.^21^ The American Academy of Pediatrics recommends introducing solid foods at approximately 6 months of age.^24^ However, data from the national longitudinal Feeding Practices Study II (2005-2007) found that approximately 40% of U.S. mothers introduced solids before 4 months of age.^25^

Although social factors like race, ethnicity, socioeconomic status, and maternal education influence antibiotic prescribing patterns,^26,27^ critical gaps remain in understanding risk factors specific to children under 2 years old, including early-life events like age at first illness, solid food introduction, mode of delivery (cesarean vs. vaginal), and maternal age. Our study addresses this gap by investigating longitudinal trends in antibiotic use among 1,032 children from birth through 2 years in the Center for Oral Health Research in Appalachia cohort 2 (COHRA2). Addressing modifiable risk factors could reduce unnecessary antibiotic use and its dual burden of childhood health complications and antimicrobial resistance.

## METHODS

### Cohort Selection

COHRA2 is a mother-child prospective cohort study that enrolled healthy, English-speaking, pregnant White women aged 18 years and older residing in North Central Appalachia, specifically West Virginia and Pittsburgh, Pennsylvania. Women were excluded if they did not have a singleton pregnancy, were immunocompromised, did not think they would remain in the geographic location for the duration of the study, did not have reliable telephone contact, or if the mother or child developed a serious health condition. Mothers were enrolled between 12 to 29 weeks of pregnancy. Of the 1172 mothers and children enrolled between November 2011 and February 2017, we included in this analysis the 1032 with information on antibiotic use in the first two years of life.

### Data Collection

Mothers gave written consent and provided their date of birth, education, medical insurance status, and household income and medical history at a prenatal, in-person visit. Information regarding the child’s date of birth, sex and delivery method and Area Deprivation Index were collected through an in-person visit in Pittsburgh and a questionnaire in West Virginia. The Area Deprivation Index (ADI), developed by the U.S. Health Resources and Services Administration, is a composite measure of neighborhood socioeconomic disadvantage based on 17 education, employment, housing quality, and poverty indicators from the American Community Survey.^29^ The ADI ranks neighborhoods from 1 (least deprived) to 100 (most deprived) and the national ADI rank was used in this analysis to enable comparisons across states. Since the ADI provided similar associations as household income and is less interpretable, it was not included in the statistical model.

During the follow-up period, mothers were interviewed every six months regarding their child’s feeding patterns, medication use, and illness. These interviews were conducted either via telephone or during the annual in person visits. At the Pittsburgh location only, study participants were interviewed every 2-3 months between the 6-month telephone interviews. These abbreviated telephone interviews were tailored to capture more accurate illness and medication timing. The every 2-to-3-month interviews were not completed in West Virginia due to regional logistical issues and therefore information regarding child’s feeding patterns, medication use, and illness timing were obtained from the 6-month telephone interviews and in-person visits.

### Illnesses, Antibiotic use, and Food introduction

During the 6-month and abbreviated interviews, mothers were asked to respond yes or no to the following question “Since we last spoke <on the phone> has <baby’s name> been sick with …. <cold, flu, ear infection, asthma, diarrhea, eczema, other>”. If the response was yes, they were asked if the child was seen by a doctor or nurse or went to hospital, and if they received a prescription for an over-the-counter medication, an antibiotic, or a medication that was not an antibiotic. Mothers were also asked to respond yes or no to the questions “During the last week did you give <baby> and infant foods or table foods?” We imputed the age of the child at time of illness, antibiotic use and food introduction as the point halfway between the interview where these characteristics were reported and the previous time of contact. For this analysis we used the imputed time of first illness, first ear infection and first introduction of solid foods.

### Statistical Analysis

We described the frequency and proportions of ever antibiotic use by study site, demographics and health characteristics using simple statistics, and tested differences in proportions by characteristic using Chi-Square tests. For time-dependent exposures, we estimated the median age at first solid food introduction and first illness using the crude median age of each event in the Pittsburgh site, where timing data were more granular.

We visualized time to first solid food introduction, time to first antibiotic use, and time to first ear infection using a cumulative incidence survival curve (the non-parametric PROC LIFETEST Kaplan-Meier estimator), right censored for loss to follow-up. To estimate hazard ratios (HR) and 95% confidence intervals, we conducted a multivariable Cox proportional hazard regression analysis predicting time to first antibiotic use. Covariates for the model were selected based on prior evidence of their association with antibiotic use. The final model included household income and birth quarter as fixed variables, and first ear infection, first solids, and first illness as time varying.

P-values < 0.05 were considered statistically significant. Analyses were conducted in SAS 9.4 (SAS Institute, Inc., Cary, NC) and RStudio Version 4.5.1. All Figures were made in RStudio Version 4.5.1.

### Ethics approval

The study protocol was approved by the Institutional Review Boards at the University of Pittsburgh (coordinating center FWA00006790) and West Virginia University (FWA00005078).^28^

## RESULTS

### Characteristics of the Study Population and Antibiotic Use

“Ever antibiotic use” within the first 24 months of life varied little by child sex, mode of delivery, and medical insurance (Table 1). However, Pittsburgh children whose mothers had higher household income (44.0% vs. 15.8%) or higher education (63.2% vs. 52.7%) were significantly more likely than children without those characteristics to ever have used antibiotics (p-value < 0.05). Conversely, in West Virginia, these relationships were not statistically significant.

**Table 1.** Ever antibiotic use among children less than 24 month by selected demographic and health characteristics, and median age at time of selected characteristics (Pittsburgh participants only). Participants in the Center for Oral Health in Appalachia 2 (COHRA2) (West Virginia N=452, Pittsburgh N=580).

|  | Ever Antibiotic Use (%) |  |  |  |  |  |
| --- | --- | --- | --- | --- | --- | --- |
|  | West Virginia | Pittsburgh | Median Age in Months of First Occurrence of Selected Characteristics - Pittsburgh |  |  |  |
| Characteristic | N<sup>a</sup> (%) | N<sup>a</sup> (%) | Antibiotic | Illness | Ear Infection | Solid Food |
| Maternal age at birth (years) |  |  |  |  |  |  |
| 18-24 | 143 (60.1) | 70 (58.6) | 8.8 | 1.7 | 8.8 | 3.1 |
| 25-29 | 129 (55.0) | 171 (58.5) | 6.6 | 2.5 | 8.6 | 4.7 |
| 30-34 | 125 (60.0) | 235 (63.8) | 8.2 | 2.6 | 8.9 | 4.8 |
| ≥ 35 | 55 (50.9) | 104 (53.8) | 9.8 | 2.5 | 11.9 | 4.7 |
| Has medical insurance<sup>b</sup> |  |  |  |  |  |  |
| Yes | 439 (57.9) | 568 (60.6) | 7.9 | 2.5 | 9.0 | 4.7 |
| No | 10 (60.0) | 9 (33.3) | 17.5 | 5.6 | 12.5 | 5.2 |
| Household income<sup>c, d</sup> |  |  |  |  |  |  |
| ≤ 50,000 | 220 (33.6) | 163 (15.8) | 8.5 | 2.5 | 9.5 | 4.5 |
| > 50,000 | 170 (24.1) | 357 (44.0) | 8.0 | 2.5 | 9.2 | 4.8 |
| Maternal education<sup>c</sup> |  |  |  |  |  |  |
| Less than college | 234 (59.4) | 186 (52.7) | 7.3 | 2.5 | 8.4 | 3.5 |
| ≥ College degree | 218 (55.5) | 394 (63.2) | 8.4 | 2.5 | 9.4 | 4.9 |
| Child's mode of delivery<sup>e</sup> |  |  |  |  |  |  |
| Vaginal | 307 (56.7) | 432 (60.0) | 7.8 | 2.5 | 9.0 | 4.7 |
| Cesarean Section | 144 (59.0) | 143 (59.4) | 8.4 | 2.4 | 9.5 | 4.6 |
| Birth quarter<sup>c</sup> |  |  |  |  |  |  |
| 01: Jan – Mar | 105 (71.4) | 138 (55.8) | 9.0 | 2.6 | 9.5 | 4.7 |
| 02: Apr – Jun | 108 (53.7) | 139 (51.8) | 7.7 | 2.5 | 9.3 | 4.7 |
| 03: Jul – Sep | 130 (45.4) | 152 (64.5) | 6.7 | 2.5 | 7.7 | 4.6 |
| 04: Oct – Dec | 109 (62.4) | 151 (66.2) | 8.4 | 2.5 | 9.2 | 4.7 |
| Child's sex |  |  |  |  |  |  |
| Male | 243 (58.4) | 307 (60.3) | 7.9 | 2.5 | 9.0 | 4.6 |
| Female | 209 (56.5) | 273 (59.3) | 8.0 | 2.5 | 9.2 | 4.7 |
| National Area Deprivation Index<sup>f</sup> |  |  |  |  |  |  |
| 4-55 (least deprived) | 113 (51.3) | 240 (64.2) | 7.7 | 2.6 | 8.7 | 4.8 |
| 56-78 | 167 (60.5) | 179 (57.5) | 8.5 | 2.5 | 8.8 | 4.7 |
| 79-100 (most deprived) | 150 (60.0) | 160 (55.6) | 8.1 | 2.5 | 9.5 | 4.4 |

<sup>a</sup> Number with characteristic, percent every antibiotic use. Percents may not add up to 100% due to missing values and rounding
<sup>b</sup> Pittsburgh data missing for 3 participants, West Virginia data missing for 4 participants
<sup>c</sup> The proportion of Pittsburgh participants with ever antibiotic use was significantly different ( $p < 0.05$ ) for these variables.
<sup>d</sup> Pittsburgh data missing for 60 participants, West Virginia data missing for 62 participants
<sup>e</sup> Pittsburgh data missing for 5 participants, West Virginia data missing for 1 participant
<sup>f</sup> Pittsburgh data missing for 1 participant, West Virginia data missing for 22 participants

The median time of first introduction to solid foods was around 4 months in both study sites. Children of younger mothers and mothers with less than a college degree were more likely to be introduced to solid foods before 4 months (Table 1). We do not have good information on breastfeeding, which is associated with later introduction of solid foods.

In both study sites, colds and ear infections were the most reported illnesses (Table 2). Antibiotic treatments for ear infections were nearly identical between West Virginia and Pittsburgh cohorts (94.1% and 94.6%, respectively). However, West Virginia children with colds were significantly more likely than Pittsburgh children to be treated with antibiotics (13.6% versus 3.3% in Year 1 and 14.1% versus 2.5% in Year 2). Notably, ear infections accounted for more than half of all reported antibiotic treatments in both cohorts—58.1% in West Virginia and 58.5% in Pittsburgh.

**Table 2.** Number of Self-Reported Illnesses and Antibiotics Prescribed by Indication, During the first 24 Months of Life^a^. Participants in the Center for Oral Health in Appalachia 2 (COHRA2) (West Virginia N=452, Pittsburgh N=580).

| Self-reported Illness | West Virginia |  |  | Pittsburgh |  |  |
| --- | --- | --- | --- | --- | --- | --- |
|  | N | Treated with Antibiotics | Percent | N | Treated with Antibiotics | Percent |
| Cold | 361 | 49 | (13.6) | 1073 | 35 | (3.3) |
| Ear Infection | 137 | 128 | (93.4) | 260 | 244 | (93.8) |
| Flu | 29 | 2 | (6.9) | 47 | 1 | (2.1) |
| Asthma | 8 | 0 | (0) | 8 | 0 | (0) |
| Diarrhea | 50 | 3 | (6.0) | 274 | 1 | (0.4) |
| Eczema | 73 | 0 | (0) | 300 | 2 | (0.7) |
| Unspecified Other | 322 | 56 | (17.4) | 902 | 142 | (15.7) |
| **Total Year 1** | 980 | 238 | (24.3) | 2864 | 425 | (14.8) |
| Cold | 269 | 38 | (14.1) | 606 | 15 | (2.5) |
| Ear Infection | 149 | 141 | (94.6) | 220 | 210 | (95.5) |
| Flu | 20 | 2 | (10.0) | 53 | 1 | (1.9) |
| Asthma | 11 | 0 | (0) | 15 | 0 | (0) |
| Diarrhea | 33 | 0 | (0) | 173 | 0 | (0) |
| Eczema | 40 | 1 | (2.5) | 166 | 2 | (1.2) |
| Unspecified Other | 116 | 37 | (31.9) | 469 | 129 | (27.5) |
| **Total Year 2** | 638 | 219 | (34.3) | 1702 | 357 | (21.0) |
| **Grand Total** | 1618 | 457 | (28.2) | 4566 | 782 | (17.1) |

<sup>a</sup>Study participants appear multiple times

### Timing of First Antibiotic Use and Illness - Cumulative Incidence of First Events

At both study sites, the median age for experiencing a first illness was around 2 months. Trends suggest that first antibiotic use and first ear infection incidence are closely related (Figure 1). For the Pittsburgh children, the median age of first antibiotic preceded the median age of first ear infection by 6 months. For West Virginia children, it appears that the median age for first antibiotic use (7 months) was nearly identical to the median age for a first ear infection (7.3 months). However, this does not reflect the true timing: telephone interviews at 2-3 months intervals were not conducted at the West Virginia site and time of event was imputed as halfway between the interviews (see Methods).

**Figure 1.**
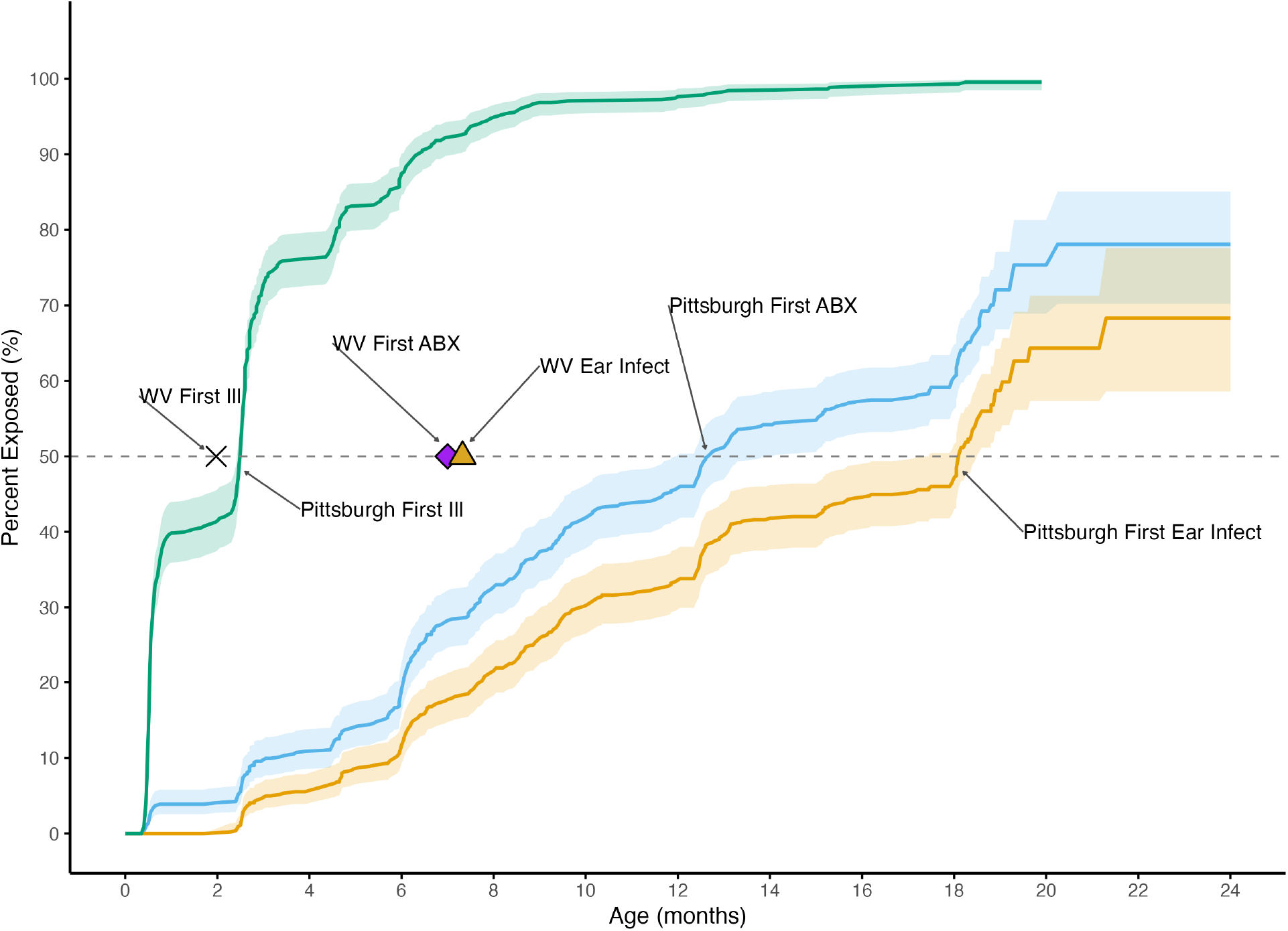
Kaplan-Meier curves showing cumulative incidence of first solid foods, antibiotic use, and ear infections in Pittsburgh children (N=452). Shaded areas represent 95% confidence intervals. Median ages for West Virginia children (N=580) are marked (X = solids; diamond = antibiotics; triangle = ear infections). Center for Oral Health in Appalachia 2.

### Factors Associated with Earlier Time to First Antibiotic Use - Survival Analysis

This analysis was limited to Pittsburgh children (see Methods). In a Cox proportional hazards model predicting time of first antibiotic use, the hazards of earlier introduction of solid food (HR; 4.75; 95% CI: 3.19, .07), earlier time of first illness (HR: 4.6; 95% CI: 3.32, 6.36); and earlier time of first ear infection (HR: 3.28; 95% CI:3.28, 4.17) were significantly associated with time of first antibiotic use by age 24 months. By contrast, there was no significant association of birth quarter or household income with time to first antibiotic use (Table 3).

**Table 3.** Results of Cox Proportional Hazard Model Identifying Associations Between Time to First Food Introduction, Time to First Illness and Time to First Ear Infection, Household Income, Birth Quarter and Time to First Antibiotic Use. Pittsburgh participants in the Center for Oral Health in Appalachia 2 (COHRA2) followed from birth to age 2 (N=580).

| Covariate | Hazard Ratio | Hazard Ratio<br>95% CI | p-value |
| --- | --- | --- | --- |
| Time of First Food Introduction |  |  |  |
| Yes | 4.75 | (3.19 - 7.07) | <.0001 |
| No | referent |  |  |
| Time of First Illness <sup>a</sup> |  |  |  |
| Yes | 4.6 | (3.32 - 6.36) | <.0001 |
| No | referent |  |  |
| Time of First Ear Infection |  |  |  |
| Yes | 3.28 | (2.58 - 4.17) | <.0001 |
| No | referent |  |  |
| Household Income |  |  |  |
| Income > 50,000 | 1.11 | (0.86 - 1.43) | 0.42 |
| Income <= 50,000 | referent |  |  |
| Birth Quarter |  |  |  |
| Quarter 2: April - June | 0.78 | (0.56 - 1.09) | 0.15 |
| Quarter 3: July - September | 1.12 | (0.82 - 1.53) | 0.49 |
| Quarter 4: October - December | 1.04 | (0.76 - 1.42) | 0.82 |
| Quarter 1: Jan - March | referent |  |  |

<sup>a</sup>First Illness includes first ear infection

### COMMENT

In this analysis of 1,032 West Appalachian children less than 24 months, we identified a novel association between timing of introduction of solid food and earlier antibiotic use. After adjustment for time of first ear infection, time of first non-ear infection, household income, and birth quarter, early introduction of solid food increased risk of first antibiotic use almost five-fold (HR: 4.75; 95% CI: 3.19–7.07). Consistent with prior evidence that ear infections and respiratory illnesses are primary drivers of pediatric antibiotic prescribing,^9^ our analysis shows that time to first antibiotic use was strongly associated with time to first ear infection and time to first illness.

In the Pittsburgh site, 78% of children were exposed to their first antibiotic by age two, a proportion higher than the 70% reported by a Minnesota study of 14,572 children and the 58% reported by a National Patient-Centered Clinical Research Network study of 1,792,849 children.^3,30^ By one year of age, 45.8% had received their first antibiotic, similar to the 40.3% reported in a Danish nationwide registry study.^31^ However, the median age at first antibiotic exposure in our cohort was approximately 12 months, younger than the 18 months reported in a 2019 study of 3,401 children in Burkina Faso.^32^

The American Academy recommends introducing solid foods at 6 months of age, but previous studies from the United States,^25^ Australia,^33^ and the United Kingdom^34^ found, as we did here, that the median age of solid food introduction was around 4 months. In these studies, reasons for introduction most often related to infant behavior, e.g., lack of sleep, rather than signs of food readiness. These studies also noted that food tended to be introduced later to infants that were still breastfeeding. We identified early solid food introduction as a strong predictor of earlier antibiotic use, but our data did not enable assessment of the modifying effect of breastfeeding on this relationship: breastfeeding is associated with reduced infection risk and therefore antibiotic use during the first 2 years of life.^35,36^ In a French national birth cohort of 10,349 children, any breastfeeding and longer duration of breastfeeding were associated with reduced antibiotic use before age two.^36^ A systematic review that included 145 primary studies and 29 systematic reviews published between 2006–2014 found that breastfeeding reduced the risk of otitis media and moderate-to-severe respiratory and gastrointestinal infections.^35^ Future studies should integrate electronic health records to validate caregiver reports and examine mediating factors such as household hygiene and daycare attendance. Additionally, studies should compare the timing of solid food introduction during versus after breastfeeding cessation to clarify their combined influence on early antibiotic exposure.

Acute otitis media (AOM), a condition whose incidence is reduced by influenza and the pneumococcal conjugate (PCV13) vaccination, remains the primary reason for pediatric antibiotic prescribing.^37,38^ Antibiotic use for AOM can be reduced by conservative treatment and monitoring. The AAP recommends antibiotics for cases that do not improve within 48 to 72 hours.^16^ A review of randomized control trials in 13 high-income countries found that approximately 60% of otitis media cases resolve within 24 hours, regardless of antibiotic use.^39^ Judicious antibiotic use is essential for slowing the emergence and spread of antibiotic resistance^40,41^; unfortunately, this goal is also challenged by inappropriate prescribing for viral illnesses. In our cohort, first illness, primarily colds, was also significantly associated with earlier first antibiotic use. Ours is not the only such report: a 2020 national point prevalence survey of 32 children’s hospitals across the U.S. (2016–2017) found that 33.4% children aged 0-17 years with an upper respiratory tract infection received antibiotics.^42^

Our study strengths include the large sample size, longitudinal design, granular follow-up data (particularly at the Pittsburgh site), and harmonized metrics across two distinct geographic sites. Although follow-up interviews were less frequent for West Virginia participants, associations observed were consistent with that observed in the Pittsburgh cohort. Our study does have some limitations. We did not have access to medical records and therefore were unable to confirm parent reports of diagnoses, prescriptions, or exact timing of illness, and we did not have sufficient data on breastfeeding duration to assess its potential modifying effect on the association with age at solid food introduction. Finally, our study population was limited to White women residing in Northern Appalachia, which may restrict generalizability to more diverse populations. Distinct sociocultural, economic, and healthcare access factors in this region may uniquely influence health outcomes, potentially limiting broader applicability of the findings.

## CONCLUSION

Antibiotic use remains pervasive in early childhood, driven by both clinical and modifiable factors. Early solid food introduction independently predicted earlier antibiotic exposure, revealing a potential avenue for prevention; however, the impact of breastfeeding within this pathway requires further study. The timing of childhood illnesses, especially ear infections, closely parallels initial antibiotic use. Risk of ear infections can be reduced by influenza and *S. pneumoniae* vaccinations and watchful waiting can limit unnecessary antibiotic use. Addressing these modifiable risk factors may help reduce unnecessary antibiotic exposure in young children.

## Acknowledgements

The authors thank COHRA2 participants and the COHRA2 study teams at the Universities of Pittsburgh and West Virginia University. Without their participation and support this student would not have been possible.

## Data Availability

The COHRA2 data (phenotypes, demographics, health history and other data) utilized in this study are available through FaceBase.org (**COHRA2**: Record ID: 4Y-4SK4; Accession: FB00001366; DOI: 10.25550/4Y-4SK4).

